# Maternal Mortality in Goiás: Causes of Maternal Death and Barriers to Underreporting

**DOI:** 10.1101/2025.03.25.25324647

**Authors:** Emily Nayana Nasmar de Melo, Otaliba Libânio de Morais Neto, Ana Maria Nogales Vasconcelos

**Affiliations:** Federal Institute of Education, Science and Technology of Goiás, Águas Lindas Campus, Águas Lindas de Goiás, Goiás, Brazil; Institute of Tropical Pathology and Public Health, Federal University of Goiás, Goiânia, Goiás, Brazil; Department of Statistics, University of Brasília, Brasília, Distrito Federal, Brazil

**Keywords:** Maternal Mortality, Information Systems, Underreporting, Cause of Death

## Abstract

**Objective:** To describe the causes of maternal deaths and estimate the magnitude of maternal mortality in the state of Goiás, considering the underreporting of maternal deaths from 2016 to 2021.

**Methods:** This is a descriptive study that linked data from the Mortality Information System (SIM in Portuguese) and the Live Birth Information System (SINASC in Portuguese) using a linkage procedure. The aim was to identify maternal deaths and their causes and calculate the Maternal Mortality Ratio (MMR).

**Results:** A total of 417 maternal deaths were officially reported in Goiás from 2016 to 2021. Among these, 291 matches were identified between the systems, including 239 cases where the declared underlying cause was maternal death, 17 with presumed causes, and 35 with other causes. The leading causes of death for maternal deaths, deaths with presumed causes, and other deaths were, respectively: other obstetric conditions not elsewhere classified; symptoms, signs, and abnormal clinical and laboratory findings not elsewhere classified; and certain infectious and parasitic diseases. Considering only declared maternal deaths, the MMR ranged from 55.79 (2016) to 137.46 (2021) deaths per 100,000 live births (LB), reflecting a 146% increase. Considering the inclusion of deaths identified through linkage, the MMR ranged from 55.79 (2016) to 157.41 (2021) deaths per 100,000 LB, representing an 182% increase.

**Conclusions:** The underestimation of MMR values was adjusted compared to direct estimates using the SIM and SINASC systems. This indicates that maternal mortality is higher than reflected in official statistics.

## INTRODUCTION

Maternal mortality remains a public health issue and a challenge for health care services. Global estimates range from 12 to 430 deaths per 100,000 live births (LB). The highest rates are recorded in Sub-Saharan Africa, South Asia, and parts of the Middle East^1’2^. It is a health indicator that reflects the quality of health care provided to the population and serves as a measure of inequalities, as it is predominantly higher in underdeveloped regions and lower socioeconomic areas of developed regions^3’4^.

In Brazil, the Maternal Mortality Ratio (MMR) ranged from 64.4 maternal deaths per 100,000 live births (LB) in 2016 to 57.9 maternal deaths per 100,000 LB in 2019. These values are very similar to those observed in the Central-West region of Brazil5. State statistics indicate that in Goiás, during the same period, the MMR ranged from 55.46 to 69.71 maternal deaths per 100,000 LB6.

This scenario underscores the reduction of maternal deaths as a key priority in public policies and global commitments, such as the Millennium Development Goals (MDGs), which aimed to reduce maternal mortality by 75% by 2015. However, the global target was not achieved7. The commitments made in 2000 were therefore extended in 2015 through the Sustainable Development Goals (SDGs). By 2030, it is expected that global maternal mortality will reach a maximum of 70 deaths per 100,000 live births, and for Brazil, a signatory of the SDGs, the target is 30 deaths per 100,000 live births8. To meet this goal, countries must achieve an annual reduction in maternal mortality of at least 6.1%, a target currently met by only 16 countries^9^.

In addition to the challenge of reducing maternal deaths, many countries still struggle with unreliable estimates of this event. Questionable vital registration and statistical systems are common, leading to incomplete or incorrect information about the causes of death^10^, obscuring the true cause and hindering its identification as maternal^11^. This is especially true in countries with high maternal mortality, such as India, which has experienced low coverage of vital records and death registrations^12-14^.

Thus, investment in actions aimed at reducing maternal mortality depends on the availability of accurate and up-to-date data^15'16^, which supports health managers and professionals’ decision-making and ensures the effectiveness and efficiency of health care services^17^. Therefore, the collected information must be accurate and relevant, to avoid misinterpretations of the population’s health^18^. In this context, improvements in measurement and standardized approaches are needed10, and it is important to identify the main causes of maternal deaths and the factors contributing to these events^19^.

Alternative approaches for investigating the true magnitude of maternal mortality have been proposed to mitigate the consequences of underreporting maternal deaths in vital statistics systems. Linkage techniques can also help verify information that has not been properly recorded^20^, as they can monitor events of interest by combining different databases, thereby providing greater quantity and quality of information on these events^21'’22^.

Considering the impact of maternal death causes on health management actions and recognizing that the availability of reliable information enables managers to take evidence-based decisions, this study aimed to describe the causes of maternal death and estimate the magnitude of maternal mortality in the state of Goiás, taking into account the underreporting of maternal deaths, from 2016 to 2021.

## METHODS

### Study type, data source, and location

This is a descriptive study in which the Mortality Information System (*Sistema de Informação sobre Mortalidade* - SIM) and the Live Birth Information System (*Sistema de Informação sobre Nascidos Vivos* - SINASC) databases were connected through the linkage procedure.

The databases included individuals’ personal information, such as name, sex, age, among others, to enable the linkage process. The SINASC and SIM databases were provided by the State Department of Health in Goiás.

The study was conducted in Goiás, a state located in the Central-West region of Brazil. It has 246 municipalities and is the 7th largest state in the country in terms of land area, covering almost the entire Federal District (the capital of Brazil)^23^.

### Linkage methodology

To facilitate the linkage process, the SIM and SINASC databases were first cleaned and organized. In the SIM database, deaths occurring from 2016 to 2021 among Women of Reproductive Age (WRA) – 10 to 49 years old – residing in Goiás were selected. Moreover, duplicate cases were excluded. In the SINASC database, mothers residing in Goiás who gave birth between 2016 and 2021 were selected, excluding duplicate and multiple birth cases. In cases where two non-multiple births were identified in the same year, the most recent birth was retained in the database.

Afterward, the SIM and SINASC databases were imported in dbf format into OpenRecLink version 3.1 for probabilistic linkage. This software is an enhanced version of Reclink, also open-source, and uses a probabilistic record linkage technique^24^.

The probabilistic method was used due to the absence of a unique identifier in the databases. By using common fields, such as name and age, automated routines calculate the probability that records from different databases correspond to the same individual^21’22^.

The linkage followed these steps: *i*. standardizing the variables (Table 1) used in the blocking step of the databases; *ii*. probabilistic matching between the databases; *iii*. generating the matched pairs file; *iv*. selecting true pairs based on the criteria of similarity in name, age, city, and address. True pairs where there was no temporal proximity of up to 42 days between the date of birth and the date of death were excluded from the matched pairs file.

**Table 1.**
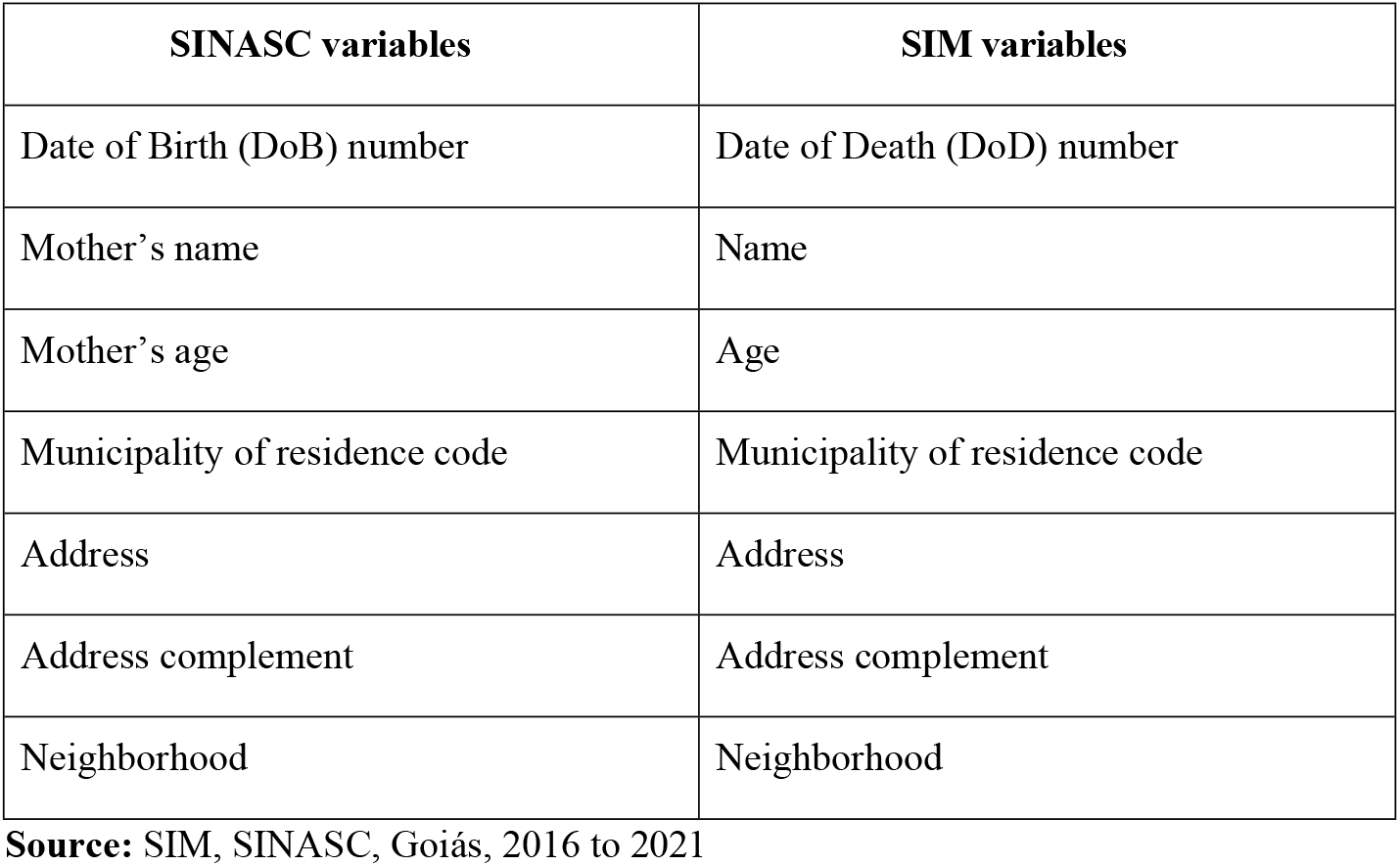
Selected Variables for Standardization and Blocking.

| SINASC variables | SIM variables |
| --- | --- |
| Date of Birth (DoB) number | Date of Death (DoD) number |
| Mother's name | Name |
| Mother's age | Age |
| Municipality of residence code | Municipality of residence code |
| Address | Address |
| Address complement | Address complement |
| Neighborhood | Neighborhood |

**Source:** SIM, SINASC, Goiás, 2016 to 2021

**Table 1.**
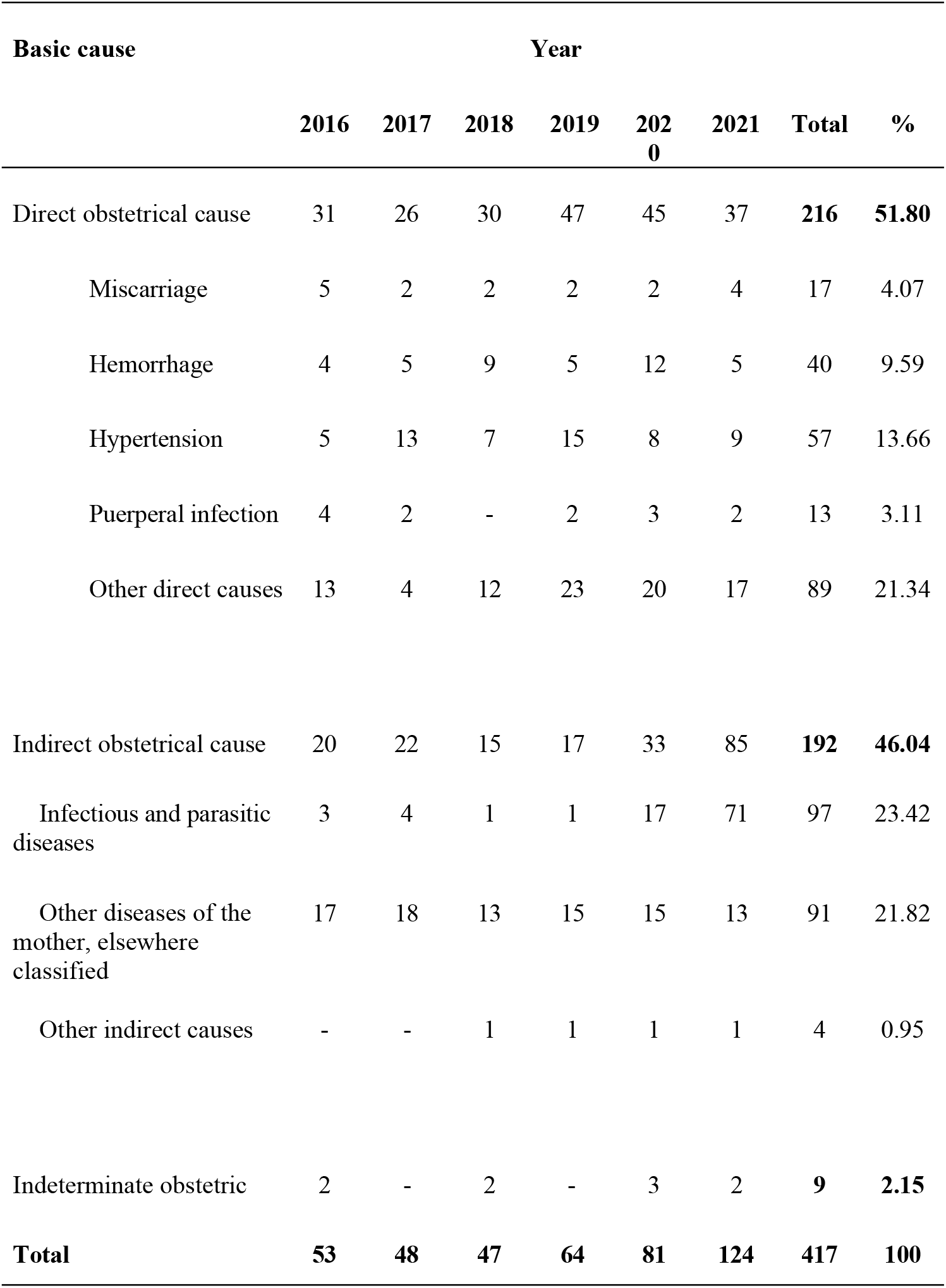
Underlying Cause of Maternal Deaths Declared, Goiás, Brazil, 2016-2021.

| Basic cause | Year |  |  |  |  |  | Total | % |
| --- | --- | --- | --- | --- | --- | --- | --- | --- |
|  | 2016 | 2017 | 2018 | 2019 | 2020 | 2021 |  |  |
| Direct obstetrical cause | 31 | 26 | 30 | 47 | 45 | 37 | **216** | **51.80** |
| Miscarriage | 5 | 2 | 2 | 2 | 2 | 4 | 17 | 4.07 |
| Hemorrhage | 4 | 5 | 9 | 5 | 12 | 5 | 40 | 9.59 |
| Hypertension | 5 | 13 | 7 | 15 | 8 | 9 | 57 | 13.66 |
| Puerperal infection | 4 | 2 | - | 2 | 3 | 2 | 13 | 3.11 |
| Other direct causes | 13 | 4 | 12 | 23 | 20 | 17 | 89 | 21.34 |
| Indirect obstetrical cause | 20 | 22 | 15 | 17 | 33 | 85 | **192** | **46.04** |
| Infectious and parasitic diseases | 3 | 4 | 1 | 1 | 17 | 71 | 97 | 23.42 |
| Other diseases of the mother, elsewhere classified | 17 | 18 | 13 | 15 | 15 | 13 | 91 | 21.82 |
| Other indirect causes | - | - | 1 | 1 | 1 | 1 | 4 | 0.95 |
| Indeterminate obstetric | 2 | - | 2 | - | 3 | 2 | **9** | **2.15** |
| **Total** | **53** | **48** | **47** | **64** | **81** | **124** | **417** | **100** |

### Database construction

The World Health Organization^25^ defines maternal death as “the death of a woman while pregnant or within 42 days of termination of pregnancy, irrespective of the duration and the site of the pregnancy, from any cause related to or aggravated by the pregnancy or its management, but not from accidental or incidental causes.” It corresponds to deaths coded in the International Classification of Diseases (ICD-10)^26^ under Chapter XV (Pregnancy, childbirth, and the puerperium) and includes the following ICD codes: A34 (Obstetrical tetanus), D39.2 (Neoplasm of uncertain or unknown behavior of the placenta), E23.0 (Hypopituitarism), F53 (Mental and behavioral disorders associated with the puerperium), and M83.0 (Puerperal osteomalacia).

Thus, deaths classified as maternal mortality were selected from the SIM database, except for those with ICD codes “O96” and “O97,” which refer, respectively, to “death from any obstetric cause occurring more than 42 days but less than one year after delivery” (late) and “death from sequelae of obstetric causes”^27^. Therefore, the second file was generated to create the database.

The files resulting from the linkage and selection of maternal deaths in the SIM database were merged, and duplicates were removed. According to the ICD-10 code registered as the underlying cause of death, the cases were grouped as follows: maternal deaths; deaths with a presumed maternal cause based on the classification of the Ministry of Health^28^; and other causes.

When a maternal death is not properly declared, meaning that instead of being associated with the gravid-puerperal period, its underlying cause is recorded as related to intermediate or terminal complications, it is classified as a death with a presumed maternal cause. The Ministry of Health^30^ provides a list of the most common “masking” causes for maternal death (A400-A403; A408-A419; A542; D65; G400-G409; G932; I10; I210-I214; I219; I269; I429; I469; I500; I509; I64; I740-I749; J100-J101; J108; J110-J111; J118; J120-J122; J128-J129; J13-J14; J150-J160; J180-J182; J188-J189; J81; K650; K658-K659; K720; N170-N172; N178-N179; N710-N711; N719; N733-N739; R568; R571; R578; R58; R98; R99; Y480-485; Y579), in addition to the material provided to the coders for causes in the SIM, which helps to select the correct underlying cause^28^.

### Data analysis

The information corresponding to Block V of the Death Certificate, which corresponds to the conditions and causes of death5, was analyzed. For the descriptive analysis of the data, SPSS version 25 was used. The MMR was calculated in two ways: i) considering the numerator consisting only of the declared maternal deaths and ii) considering the numerator formed by the combination of declared maternal deaths and deaths identified through linkage, excluding those caused by accidents or incidental causes. The MMR calculation is defined by dividing the number of maternal deaths by the number of live births during the same period, then multiplying by 100,000 (MS, 2009). The MMR was calculated with its respective confidence interval.

### Ethical considerations

This study was conducted in accordance with the principles of Resolutions 466/2012^31^ and 580/2018^32^. It was approved by the Research Ethics Committee of the Federal University of Goiás, under approval 5.261.335, and has the consent of the State Health Department for the provision of the nominal databases.

## RESULTS

From 2016 to 2021, 417 maternal deaths were recorded in Goiás in the SIM, resulting in 44 fetal deaths and 17 abortions. The linkage process identified 291 pairs between the SIM and SINASC, of which the underlying cause of 239 cases was maternal death, 17 were classified as presumed maternal death, and 35 had other causes, as shown in Figure 1. Thus, 67.1% of the Death Certificates of women listed as mothers in the Live Birth Declarations (*Declaração de Nascidos Vivos* - DNV) were linked through the linkage procedure.

**Figure 1.**
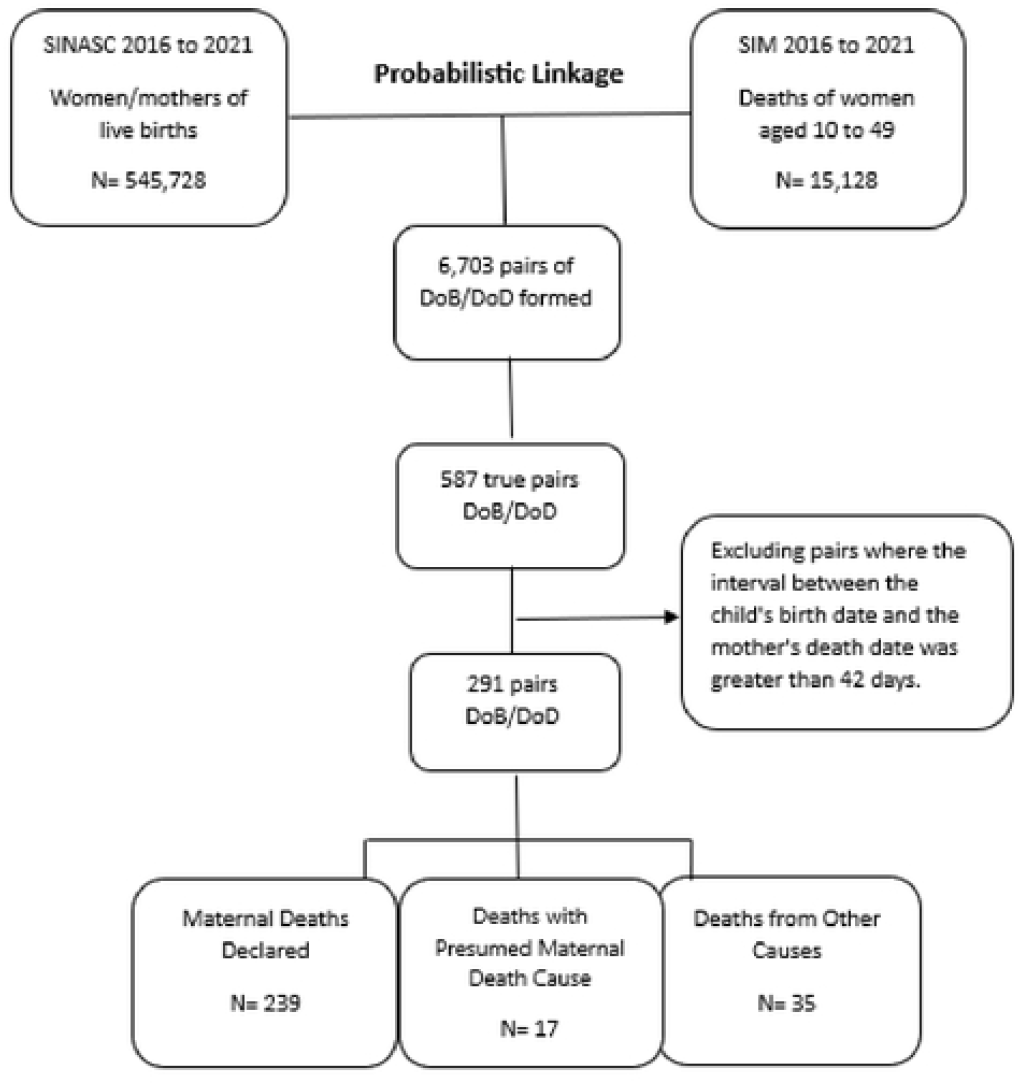
Linkage between the SIM and SINASC databases, 2016 to 2021, Goiás.

Table 1 shows the main causes of maternal deaths by year in Goiás. In the analyzed period, most of the deaths occurred due to direct obstetric causes (51.55%), mainly hypertensive diseases (13.66%), followed by hemorrhage (9.59%). In the stratification by year, only in 2021 were indirect obstetrical causes the main cause of maternal deaths, primarily due to infectious and parasitic diseases.

Among the deaths matched in the linkage that were not classified as maternal deaths, two groups were identified: one with deaths having a presumed maternal cause according to the Ministry of Health classification, and the second group consisting of deaths from other causes (Figure 2). In the group formed by presumable causes, conditions (R57.1 and R99) from Chapter XVIII (Symptoms, signs, and abnormal clinical and laboratory findings, not elsewhere classified) of the ICD-10 were the leading cause of death (52.93%) in the analyzed period. In the group with other causes of death, conditions (A90; A91; B342; B948) from Chapter I (Certain infectious and parasitic diseases) of the ICD-10 were the leading cause of maternal deaths, followed by conditions (X70.0, X70.9, X95.0, X95.4, X99.0) from Chapter XX (External causes of morbidity and mortality).

**Figure 2.**
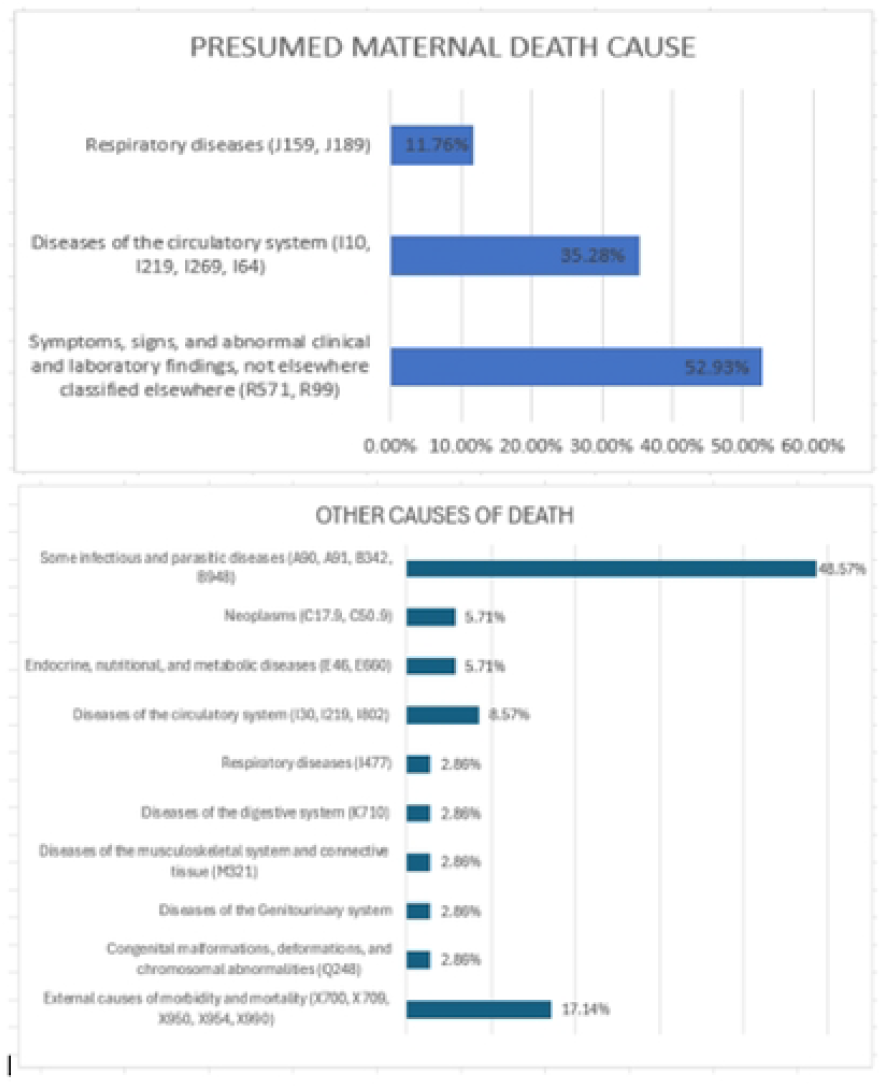
Causes of deaths identified in the linkage not classified as maternal deaths Among the deaths with ICDs in other categories, three cases were identified that presented maternal ICD in the Death Certificate listing antecedent or consequential causes, or significant conditions contributing to the death. The basic causes of death and their associated conditions were: E46 (Unspecified protein-energy malnutrition) listed with F53.0 (Mild mental and behavioral disorders associated with the puerperium, not elsewhere classified); E66.0 (Obesity due to excess calories) listed with O13 (Gestational [pregnancy-induced] hypertension); I61.9 (Unspecified intracerebral hemorrhage) listed with O15.9 (Eclampsia, unspecified as to time period).

Regarding the completion of the period in which the death occurred, most of the declared maternal deaths (60.67%) happened during the puerperal period (up to 42 days after childbirth), as well as the deaths with presumed causes (42.85%) and other causes (31.42%). Inadequate completion was identified in 2.86% of maternal deaths, where it was recorded that the death occurred outside the gravid-puerperal period. In non-maternal deaths, the information about the period in which the death occurred was ignored in most cases (Table 2). From the total maternal deaths, approximately 77.9% were investigated.

**Table 2.**
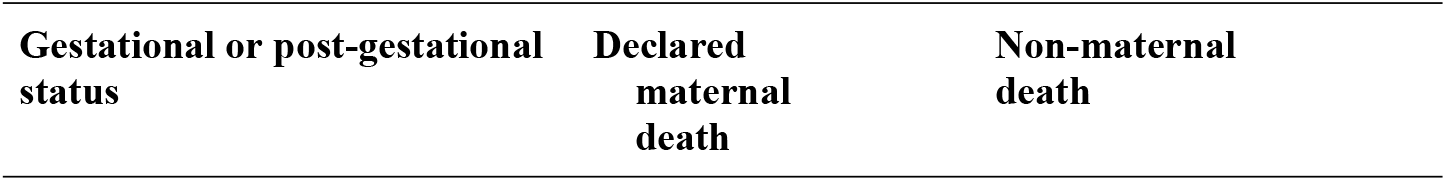
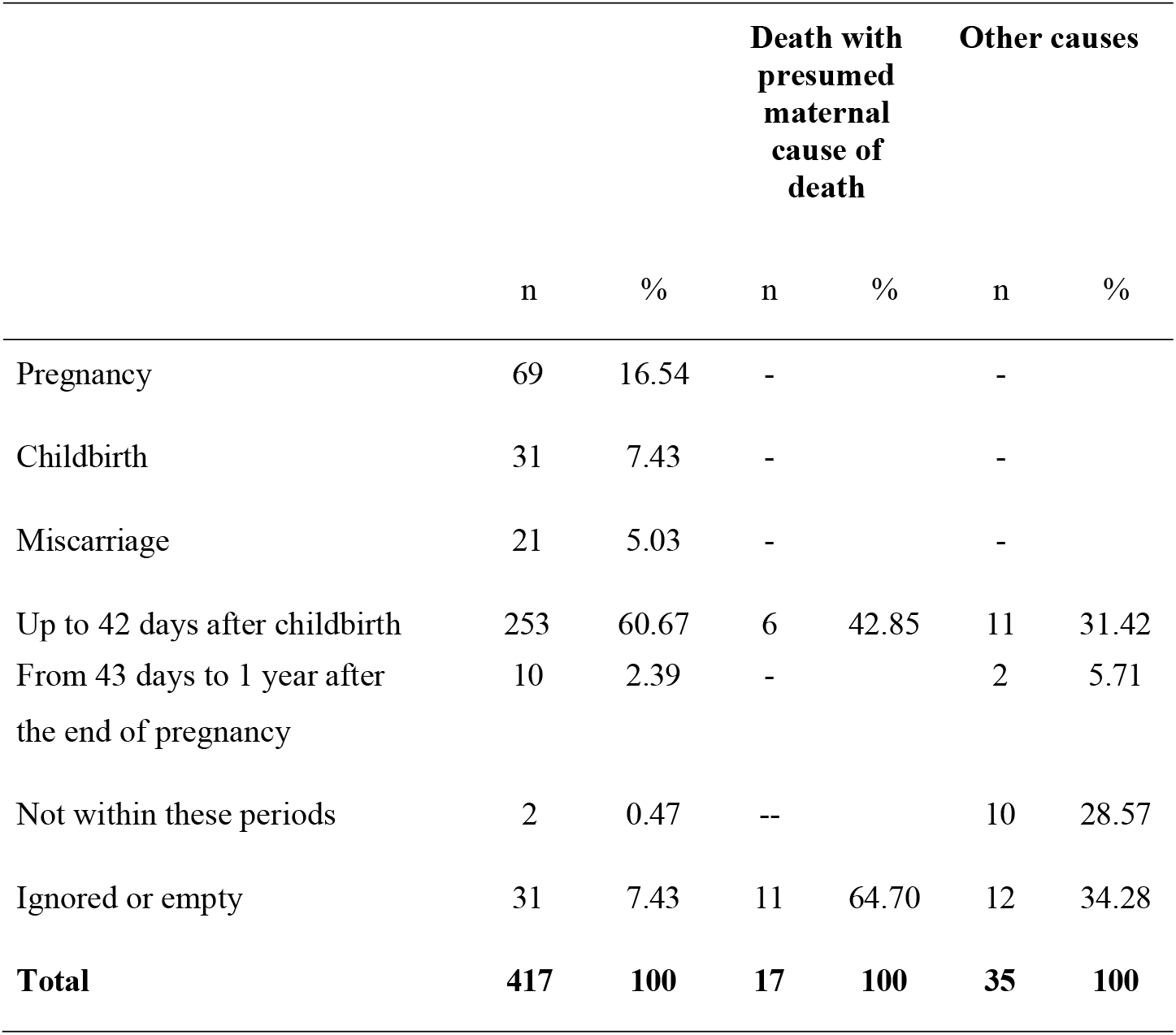
Classification of death according to the gestational or post-gestational status and the cause of death.

Considering only the declared maternal deaths, the MMR ranged from 55.79 (95% CI 40.7–70.8) deaths per 100,000 live births (LB) in 2016 to 137.46 (95% CI 113.3–161.6) maternal deaths per 100,000 LB in 2021, indicating a growth of 146.38% in the MMR over the analyzed period. After including the deaths identified in the linkage, the corrected MMR was 55.79 deaths per 100,000 LB in 2016 and 157.41 deaths per 100,000 LB in 2021 (Table 3), indicating an 182.14% increase in the corrected MMR. During the analyzed period, the MMR was corrected by 9.85% after the linkage, and in 2018, the highest increase in the number of deaths was identified after the linkage, with an adjustment in the MMR of 24.19%. There was no change in the MMR for 2016, as no additional deaths were identified through the linkage in that year.

**Table 3.** Maternal Mortality Ratio Before and After Linkage, 2016 to 2021.

| Year | LB | Declared maternal deaths | MMR (IC95%) | Maternal deaths identified in the linkage* | MMR (IC95%) | Growth |
| --- | --- | --- | --- | --- | --- | --- |
| 2016 | 94,999 | 53 | 55.79 (40.79-70.8) | 53 | 55.79 (40.78-70.8) | 0% |
| 2017 | 74,505 | 48 | 64.42 (46.21-82.64) | 50 | 67.10 (48.52-85.7) | 4.99% |
| 2018 | 98,255 | 47 | 47.83 (34.16-61.51) | 62 | 63.10 (47.4-78.8) | 24.19% |
| 2019 | 94,657 | 64 | 67.61 (51.05-84.17) | 71 | 75.00 (57.57-92.45) | 9.85 |
| 2020 | 92,107 | 81 | 87.94 (68.8-107.1) | 85 | 92.28 (72.68-111.19) | 4.70 |
| 2021 | 90,205 | 124 | 137.46 (113.3-161.6) | 142 | 157.41 (131.5-183.3) | 12.67% |
| **Total** | **544,728** | **417** | **76.55 (69.21-83.9)** | **463** | **84.99(77.26-92.73)** | **9.93%** |

\*p<0.0001

## DISCUSSION

According to the results of this study, we identified 52 deaths of pregnant or postpartum women who died within 42 days after childbirth between 2016 and 2021, in addition to the 417 maternal deaths recorded in the SIM. It can be inferred that these deaths result from incorrect classification when completing the Death Certificate, underestimating the maternal mortality rate in Goiás by approximately 10%.

In this study, 17 deaths were associated with a presumed maternal cause of death, which corroborates the findings of Gomes *et al*.^33^ for the states of São Paulo, Paraná, Pará, Ceará, and Mato Grosso, in 1999 and 2000. The authors identified 55 masked maternal deaths among 651 deaths declared by the system. In the study by Szwarcwald *et al*.^34^, approximately 1% of deaths with presumed maternal causes and other causes in Brazil were identified and reclassified, from 2009 to 2011.

A study that used the Reproductive Age Mortality Study (RAMOS) methodology to estimate the MMR in the state capitals in Brazil and the Federal District also identified 95 deaths that were classified as maternal deaths after investigation. This allowed for the correction factor for these deaths to be obtained for Brazil and its five regions^3^. The correction factor for maternal deaths is a calculation proposed to reduce the underestimation of maternal deaths, which is the ratio between the sum of maternal deaths recovered after investigations and the official declared maternal deaths, excluding those not considered as maternal deaths, divided by the latter value^35^.

After improving vital registration systems, there is likely to be an increase in the number of maternal deaths recorded with greater accuracy. Although more frequent, the underreporting of maternal deaths is not an isolated issue in low- and middle-income countries. Studies conducted in different countries, including high-income ones, show that many maternal deaths are not recorded or classified properly in information systems^36’37^. Therefore, methods that identify and investigate maternal deaths need to be improved to understand the reality and implement effective prevention and care measures.

More efficient records also indicate the main causes of maternal death, which are essential for reducing maternal mortality and making effective decisions regarding policies and programs^38^. In our study, most deaths occurred due to direct obstetric causes (51.55%), primarily from hypertensive diseases and hemorrhage. International studies^39−41^ and national studies^42’43^ also report similar findings.

Regarding deaths classified under other categories, infectious and parasitic diseases can be mentioned as the leading cause of death in this classification. Many of these deaths were due to COVID-19 cases in 2020 and 2021^44^.

Although external causes are not included in the MMR calculation, cases of deaths due to violence highlighted in this study are worth noting. We identified 2 deaths from self-reported violence and 4 homicides in women who were in the gravid-puerperal period, which is a sensitive factor for maternal death notification but requires attention and further research investment. Although homicide and suicide cases during the gravid-puerperal period are rare, and less emphasis is placed on detecting and preventing them, improving identification and monitoring methods could raise awareness among healthcare professionals about these deaths, which could often be prevented^45^.

In addition to underreporting, our study identified flaws in the information regarding the timing of the gravid-puerperal period in which the death occurred, with incorrect or incomplete data. Correctly classifying the death as maternal remains a challenge for health care systems^46’47^

Despite some incorrect information, it was identified that most of the deaths in this study occurred in the postpartum period (up to 42 days after delivery). This corroborates the study by Qomariyah *et al^19^*, where 55.4% of maternal deaths in three districts of the Banten province in 2016 occurred in the postpartum period.

Laurenti, Jorge, Gotlieb^3^ emphasize that the recording of pregnancy/postpartum in a variable does not directly imply a maternal death case, but rather serves as an alert for a potential maternal death, which requires detailed investigation to confirm or rule out.

Approximately 77.9% of the deaths were investigated, and every maternal death or death of a woman of reproductive age must be investigated, regardless of the reported cause. The investigation should be concluded within a maximum of 120 days from the date of occurrence^48^.

Efforts have been made to improve registration systems. For example, in the United States (USA), a 2003 update introduced an option in the Death Certificate to be marked when a death occurred during pregnancy, aiming to identify maternal deaths more easily^37^. In Switzerland, since 2007, link birth data have been linked with the mother’s date of death, thus enabling the identification of maternal deaths in women who had children up to a year prior to their death.

Several studies^34’49^ have also been developed to present alternative methods for investigating the underreporting of maternal deaths. Anwar *et al*. ^50^, who used an enhanced surveillance system, identified a 27% underestimation of maternal mortality in Pakistan.

Many of these applied methodologies corroborate the findings of our study, which show an increase in the MMR by identifying more deaths than those officially reported. We identified an increase in the MMR between 2016 and 2021, both for the declared maternal deaths and for the corrected MMR. However, a study conducted in Indonesia showed a significant reduction in maternal mortality after implementing an information monitoring system in hospitals and health units, possibly due to improving obstetric care supported by the new program^51^.

This suggests that timely monitoring of actions related to obstetric care may be the key to reducing maternal deaths. In their research on the Italian Obstetric Surveillance System, Donati *et al*.^39^ present that proactive surveillance, with systematic data collection, detailed analysis through audits and confidential surveys, and implementation of measures based on these data, contributes to promoting a culture of accountability and defining concrete and effective actions to minimize avoidable maternal morbidity and mortality.

Regarding limitations of the study, the use of secondary databases from the SIM and SINASC should be considered, which may present underreporting of total deaths and births. Not having a unique identifier between these systems may limit the identification of an individual in both systems.

This research demonstrates that the collected data enabled us to adjust the underestimation of MMR values in relation to direct estimates using SIM and SINASC, indicating that maternal mortality is higher than official statistics. The results revealed that the MMR in the state of Goiás remains high and significantly above the reduction target set for 2030.

Thus, this study highlights that many deaths of women during the gravid-puerperal period are still not properly classified as maternal deaths, and that not all deaths of women of reproductive age are investigated as recommended. Vital registration systems need to be improved for accurate quantification of maternal mortality and its primary causes, so that public policies can be refined to effectively reduce maternal mortality.

## Data Availability

We hereby declare that we are making the data used in this research partially available. We have sent a file with de-identified data containing all the information that was analyzed in our study. The part of the data that was not sent is personal information, such as full name and address, of the research participants. To also obtain access to this data, it is necessary to obtain approval from the Leide das Neves Ethics Committee and subsequent provision of the data by the Health Department of the State of Goiás, Brazil.

